# The Clinical and Economic Value of a Successful Shutdown During the SARS-CoV-2 Pandemic in Germany

**DOI:** 10.1101/2020.05.12.20098996

**Authors:** Afschin Gandjour

## Abstract

**Background and aim:** A shutdown of businesses enacted during the SARS-CoV-2 pandemic can serve different goals, e.g., preventing the intensive care unit (ICU) capacity from being overwhelmed (‘flattening the curve’) or keeping the reproduction number substantially below one (‘squashing the curve’). The aim of this study was to determine the clinical and economic value of a shutdown that is successful in ‘flattening’ or ‘squashing the curve’ in Germany.

**Methods:** In the base case, the study compared a successful shutdown to a worst-case scenario with no ICU capacity left to treat COVID-19 patients. To this end, a decision model was developed using, e.g., information on age-specific fatality rates, ICU outcomes, and the herd protection threshold. The value of an additional life year was borrowed from new, innovative oncological drugs, as cancer reflects a condition with a similar morbidity and mortality burden in the general population in the short term as COVID-19.

**Results:** A shutdown that is successful in ‘flattening the curve’ is projected to yield an average health gain between 0.02 and 0.08 life years (0.2 to 0.9 months) per capita in the German population. The corresponding economic value ranges between €1543 and €8027 per capita or, extrapolated to the total population, 4% to 19% of the gross domestic product (GDP) in 2019. A shutdown that is successful in ‘squashing the curve’ is expected to yield a minimum health gain of 0.10 life years (1.2 months) per capita, corresponding to 24% of the GDP in 2019. Results are particularly sensitive to mortality data and the prevalence of undetected cases.

**Conclusion:** A successful shutdown is forecasted to yield a considerable gain in life years in the German population. Nevertheless, questions around the affordability and underfunding of other parts of the healthcare system emerge.

## Introduction

Coronavirus disease 2019 (COVID-19) is an infectious disease caused by severe acute respiratory syndrome coronavirus 2 (SARS-CoV-2). A SARS-CoV-2 outbreak was identified first in Wuhan, Hubei, China, in December 2019. The SARS-CoV-2 epidemic was recognized as a pandemic (a worldwide spread of a new disease (WHO 2010)) by the World Health Organization (WHO) on March 11, 2020. It was confirmed to have been transmitted to Germany on January 27, 2020. As of June 15, 2020, the number of reported COVID-19 cases in Germany is 186,461, while the case fatality rate (CFR) is 4.7%. The latter may present an underestimate due to a time window between the beginning of an infection and death. On the other hand, it may present an overestimate due to undiagnosed cases in the population and deaths of COVID-19 patients attributable to concomitant diseases. The median age of death was 82 years on June 15, 2020 (Robert Koch Institut 2020).

The German federal government and the German federal states have responded with travel restrictions and the closures of schools, universities, restaurants, cafes, bars, and other public and private entities. Although some restrictions have been lifted in the meantime, others remain in place. In Germany, a major purpose of the shutdown of businesses has been to postpone the pandemic wave (‘flattening the curve’) to avoid overstretching intensive care capacity at the time of peak demand. Hence, intensive care capacity presents a critical bottleneck in responding to the pandemic. ‘Flattening the curve’ may thus buy time to expand health and intensive care capacities as well as to develop and test new vaccines, monoclonal antibodies, or drugs. The process of developing a vaccine and obtaining market approval has been estimated to take approximately 12 to 18 months (WSJ 2020). Moreover, there is a “race” to find COVID-19 treatments by repurposing drugs that are already approved for other diseases and have acceptable safety profiles (Kupferschmidt 2020). Given the necessary time lag, however, ‘flattening the curve’ may only be able to halt the epidemic by achieving herd immunity through natural infection and not through vaccination.

A more rigorous strategy than ‘flattening the curve’, which has also received notable attention, is stopping or ‘squashing’ the curve. This strategy aims at suppressing the pandemic, by bringing the reproduction number (the average number of secondary infections due to a single primary infected person) substantially below 1, until an effective vaccine or treatment becomes available.

As a result of the SARS-CoV-2 pandemic and the governmental response, the German economy is being hit by a combined supply and demand shock (Bundesministerium für Wirtschaft und Energie 2020). Trade-offs between protecting lives and revitalizing the economy thus seem inevitable. Therefore, the purpose of this study is to determine the clinical and economic value of minimizing the number of life years lost due to the SARS-CoV-2 pandemic in Germany. While I will refer specifically to a ‘shutdown’ in the following, COVID-19 containment measures go beyond a shutdown of businesses and include tracking, testing, public mask-wearing, and other measures that are largely independent of a shutdown. The study takes an ex-ante viewpoint, i.e., before a potential surge of COVID-19 cases (e.g., in a second pandemic wave) possibly overstretches ICU capacity. I determine the clinical and economic value both of ‘squashing’ and ‘flattening’ the curve, thus assuming, respectively, that a new vaccine will and will not be available before herd immunity through national infection is achieved. Hence, while ‘flattening the curve’ slows the spread of SARS-CoV-2 infections and hence prevents a sharp increase in the number of new COVID-19 cases, it is not assumed to prevent herd immunity through national infection. As the study does not analyze specific subgroups of the population but the population in aggregate, minimizing the number of life years lost by ‘flattening the curve’ thus effectively translates into minimizing lives lost as well as minimizing the CFR at the time of herd immunity. In contrast, when ‘squashing the curve’, losses of life years and lives are minimized by reducing the incidence of SARS-CoV-2 infections until the time a new vaccine will be available. Moreover, when ‘flattening the curve’ the economic value of minimizing the number of life years lost corresponds to the maximum economic value of a shutdown that is 100% effective at preventing an overstretch of ICU capacity (because a successful shutdown leads to a minimization of lives lost and life years lost). Hence, under a scenario in which the spread of the virus in the population is unavoidable, a successful shutdown presents the best-case scenario.

As a comparator of a shutdown that is successful in ‘flattening’ or ‘squashing” the curve, I considered a scenario with no ICU capacity left to treat COVID-19 patients. While this scenario presents an extreme (worst) case, it is appropriate to determine the maximum health benefit of a successful shutdown and calculate the maximum economic value of the shutdown. This allows a comparison with actual and expected public spending during and after the SARS-CoV-2 pandemic in Germany. In addition, for ‘flattening the curve’ the study considers alternative scenarios with ICU capacity being exceeded by varying amounts. These scenarios help assess the maximum economic value of freeing up or adding ICU capacity while ‘flattening the curve’.

## Methods

### Life-table model

As a basis for the calculation of life years saved by a successful shutdown and alternative scenarios, I developed a life-table model to summarize the age-specific mortality impact of the SARS-CoV-2 pandemic. In order to account for an increase in mortality due to the SARS-CoV-2 pandemic, I used two methodological approaches. In the first, I multiplied the probabilities of survival given exposure to SARS-CoV-2 with the probabilities of survival from competing causes of death. This calculation relies on an independence assumption, implying that individuals not dying from COVID 19 have the same probability of death as all individuals before the rise of the pandemic. Given that patients who die from COVID 19 tend to have more comorbidities (Wu 2020), I assumed a harvesting effect in the second approach. This approach presumes that those who die from COVID-19 are sicker and “would have died anyway” (cf. Financial Times 2020). In this scenario, I assumed for age groups with excess mortality associated with COVID-19 (the difference between observed and prepandemic mortality rates) that except for COVID-19, there are no other causes of death in the forthcoming 12 months. This is compatible with the notion that COVID-19 may be “the cause of all fatalities” (Centre for Evidence-Based Medicine 2020). It also implies that those who do not die from COVID-19 despite being infected represent a healthier population. In either approach, the population that is not infected (which is one minus the proportion of the population that has recovered from COVID-19 to provide herd immunity) is assumed to remain at risk for competing causes of death.

Assuming that deaths occur, on average, halfway at each age, I took the average of the number of people at the start and end of the age interval to estimate state membership (Barendregt 2009). To calculate life-expectancy gains of a successful shut-down and alternative scenarios, I determined the cumulative probability of an individual at age *i* of surviving until age *j* (i.e., the product of age-specific survival probabilities up to age *j*) as obtained from the life table. I took the sum over all ages *j*, thus obtaining the remaining life expectancy of an individual at age *i*. The remaining life expectancy needs to be interpreted as a hypothetical measure that summarizes the age-specific death rates in a population exposed to SARS-CoV-2. I determined the difference between a successful shutdown (and alternative scenarios) and no intervention, thus obtaining the incremental number of life years gained. To account for the age distribution of the population, I weighted age-specific life-expectancy gains by age-specific population sizes. I performed all calculations for men and women separately and then aggregated the results.

As updating the probabilities of survival in the life table and calculating the remaining life expectancy yields the remaining life expectancy (and associated loss) with life-long exposure to SARS-CoV-2 (as opposed to a one-time exposure), it was adjusted accordingly. To this end, I distributed the age-and gender-specific loss in life expectancy over the age-specific remaining lifetime before the pandemic by dividing the two variables and then aggregated the resulting figures across age and gender by the corresponding population sizes. I did not discount health benefits because the reported survival benefits from cancer treatment (see below), which are used to determine the economic value of a life year, were undiscounted as well.

### Scenario analysis

In the scenarios with insufficient ICU capacity, all patients barred from admission to the ICU were assumed to die. However, even with sufficient ICU capacity patients face a probability of death both in the ICU and post discharge. The resulting mortality is considered to be unavoidable (with currently available treatments) and, hence, cancels out in the estimate of life years associated with a shutdown that is successful in ‘flattening the curve’ versus no intervention. More specifically, I assumed that fatality in the ICU is already reflected in the CFR reported for Germany before the occurrence of the pandemic peak. Hence, I only added the fatality one year after discharge (multiplied by the proportion of the population admitted to the ICU) to the currently reported population CFR.

In addition, I considered 4 scenarios with varying degrees of insufficient ICU capacity. To determine the corresponding loss of life years compared to the situation before the pandemic, I calculated a weighted-average loss of life years per capita in each scenario, with weights reflecting the proportions of patients admitted to the ICU and refused admission. These weights were multiplied by the average loss of life years when all patients with indication for the ICU are admitted and refused admission. Of note, the excess demand for ICU beds available during the crisis refers to the average demand and not the peak demand at the height of the SARS-CoV-2 pandemic.

Finally, I analyzed a scenario based on the idea of ‘squashing the curve’. To this end, I applied the mortality data before the pandemic and calculated the difference in life years gained compared to no intervention. The calculation is equivalent to adding the absolute loss of life years associated with a shutdown that is successful in ‘flattening the curve’ to the gain in life years of the latter compared to no intervention.

### Sensitivity Analysis

In one-way deterministic analyses, I assessed parameter uncertainty by varying the input parameters that are subject to variation one at a time.

### Valuation of life years

Given the absence of an official cost-effectiveness threshold and lack of value-based prices for treatments of COVID-19 patients in Germany, which may allow deriving the willingness to pay for an effective shutdown (cf. Gandjour 2020), I borrowed the willingness to pay from the cost-effectiveness ratio of new, innovative cancer drugs. In Germany, the prices of new, innovative drugs are subject to negotiation between manufacturers and representatives of statutory health insurance (SHI). Negotiated list prices hold for all German citizens, including those covered by private insurance. From the perspective of an average citizen, notable similarities between COVID-19 and cancer exist. First, with regard to the next 12 months (the earliest point in time a vaccine is expected to become available), the expected death toll from cancer will fall in a similar range as the death toll from COVID-19 if herd immunity is achieved through natural infection (approximately 223,000 people in Germany died of cancer in 2016 (Robert Koch Institut 2016)). Hence, from the perspective of an average citizen, deaths from cancer will not be more remote than deaths from COVID-19. Therefore, at the population level, both diseases seem to pose a similar threat to life within the next 12 months. Based on this framework, beyond the 12-month period there is no additional benefit of a vaccine for COVID-19 because the maximum benefit of a vaccine is already realized in the forthcoming 12 months by avoiding the death toll from natural infection.

Moreover, from the perspective of an average citizen, the probability of being affected by a severe disease in the next 12 months is also comparable. Approximately 1.6 million Germans suffer from cancer (diagnosed within the past 5 years), while the number of COVID-19 cases expected to require ICU treatment^1^ within the next 12 months under ‘flattening the curve’ is 4.4 million (bearing in mind that this is likely to present an overestimate due to a high number of undetected cases (Streeck 2020)). In addition, for both diseases, the vast majority of people is not expected to suffer.^2^ While the time from diagnosis to death is usually longer in the case of cancer, this should increase the value of treating cancer rather than decreasing it. That is, the considerable loss in the quality of life that comes before death occurs on top of the mortality burden itself. The relevance of a direct comparison between COVID-19 and cancer for priority setting has been confirmed in two recent studies conducted in Germany and the United States (U.S.) (Brunner 2020, Sud 2020). Both studies stress the importance of avoiding delays in cancer surgery despite the need to treat hospitalized COVID-19 patients.

There are other reasons why the prices of new cancer drugs provide an upper bound for the monetary value. They have been controversially discussed in the public and are partly driven by the costs of research and development. Furthermore, the average age of death from cancer is lower (73 years (Robert Koch Institut 2016)) versus 81 years in COVID-19).

Notably, the contingent-valuation method, which asks members of a representative sample of the population for their hypothetical willingness to pay, requires educating participants about epidemiological concepts, such as excess mortality and competing diseases, which are key for understanding the mortality burden in COVID-19. That is, a survey cannot simply focus on COVID-19 and ignore concomitant diseases. However, even if this were feasible, contingent-valuation techniques, such as discrete choice experiments, have not found their way into the official drug assessment and pricing policy of pharmaceuticals in Germany, despite being tested over many years and even in a perhaps less controversial role, i.e., weighting the adverse events and desirable outcomes of drugs. Therefore, it seems unlikely that a population survey using a discrete choice experiment or an alternative contingent-valuation technique could influence policymaking in this crisis. Furthermore, politicians who are involved in current policymaking accept the negotiation outcomes for pharmaceuticals (otherwise, the negotiation process would be on the political agenda). Hence, politicians may also have a more favorable view of using the negotiation results for the purpose of putting a price tag on the life years gained from the shutdown.

Given that the economic value would need to be compared against the direct (non-)medical costs to treat COVID-19 patients as well as the indirect costs secondary to productivity loss, the perspective of the analysis is inherently societal. Given the societal perspective, I made the simplifying assumption that cancer drug costs from the SHI perspective are equal to the drug costs from a societal perspective. Strictly speaking, this does not hold particularly if manufacturers reside inside Germany. In the latter case, drug expenditures need to be adjusted for producer surplus, as it presents a gain in societal welfare (Garrison 2010).

## Data

### Health benefits

The model input data are listed in Table 1. I used the most recent period life table of the German Federal Office of Statistics (2019), which incorporates the mortality data by age and gender up to the age of 100 years (data are from 2016 to 2018). In addition, I used data from the German Federal Office of Statistics on the population size by age and gender up to the age of 100 years.

**Table 1.** Input data used in the base case and the sensitivity analysis.

| Input | Mean (range) | Reference |
| --- | --- | --- |
| Probability of death by age and gender in Germany | see reference | Federal Office of Statistics 2019 |
| Population size by age | see reference | Federal Office of Statistics 2020 |
| CFR in Germany |  | Robert Koch Institut 2020 |
| Total population | 0.047 (0.0036 – 0.047) |  |
| 0-9 years | 0.0002 |  |
| 10-19 years | 0.0002 |  |
| 20-49 years | 0.0012 |  |
| 50-69 years | 0.0197 |  |
| 70-89 years | 0.1987 |  |
| 90+ years | 0.3111 |  |
| Probability of ICU indication | 0.076 (0.04 – 0.08) | Robert Koch Institut 2020 |
| CFR in the ICU | 0.25 (0.21 – 0.52) | Robert Koch Institut 2020 |
| False-positive ICU admissions | 0.1 (0.1 – 0.2) | Abers 2014 |
| CFR one year post ICU discharge | 0.59 (0.47 – 0.73) | Damuth 2015 |
| Herd protection threshold | 0.70 (0.60 – 0.70) | Kwok 2020 |

ICU = intensive care unit, CFR = case fatality rate

Data on overall case fatality in the German population and in 6 age groups as well as data on ICU fatality were obtained from the Robert Koch Institut (2020). Data on 101 cases could not be classified by age. In a sensitivity analysis I applied the CFR from a recent empirical study on 1000 people in one German district (Heinsberg), which shows a rate of just 0.36% (Streeck 2020) and thus suggests a high number of undetected cases in the national data. I adjusted the percentage of patients admitted to the ICU accordingly because a lower CFR also implies a lower percentage of cases admitted to the ICU. The base-case admission rate to the ICU is 7.6% (Robert Koch Institut 2020). I assumed a 10% rate of inappropriate (false positive) ICU admissions in the base case. A rate above 0% seems plausible given that “good clinical practice demands that greater emphasis be placed on patient safety by limiting false negatives” (Abers 2014). This strategy comes at the risk of excessive use of ICU facilities. Nevertheless, even a rate of 10% has been considered “exceptionally low” in a non-COVID-19 setting (Abers 2014). Therefore, I increased the rate to 20% in a sensitivity analysis.

In Germany, the fatality rate in the ICU for all COVID-19 patients is 28% (Robert Koch Institut 2020). Sixty-nine percent of patients currently being treated in the ICU due to COVID-19 receive mechanical ventilation (Robert Koch Institut 2020). Given that ICU survivors face an increased probability of death after discharge, I added the difference between the ICU fatality rate and the postdischarge fatality rate from another study (Damuth 2015) to the ICU fatality rate reported by the Robert Koch Institut (2020). Data on the former source were obtained from a meta-analysis of international studies on critically ill patients treated with prolonged mechanical ventilation (Damuth 2015). In this study, the pooled mortality at hospital discharge was 29% and thus is close to the rate reported for COVID-19 ICU patients. The one-year mortality increased to 59%. Nevertheless, the meta-analysis may be criticized because the data are relatively old (studies were published before November 2013) and heterogeneous in terms of outcomes. For example, the U.S. showed a significantly higher mortality than the rest of the world. Nevertheless, the fact that the mortality at hospital discharge exactly matches the current data for COVID-19 patients seems to be a convincing argument in favor of incorporating this data from the meta-analysis. In a sensitivity analysis, I incorporated the one-year CFRs reported for the U.S. and the rest of the countries (73% and 47%, respectively).

A considerable increase in mortality after hospital discharge was confirmed in a recently published study on 21 COVID-19 patients admitted to the ICU at a single center in Washington State (Arentz 2020). In this study CFR was 52% at hospital discharge and increased to 67% 12 days after admission. In a sensitivity analysis, I incorporated the CFR on the ICU from this study as an upper bound. As the lower bound, I used data from an analysis of the European Surveillance System (TESSy) database including 13,368 patients admitted to the ICU with laboratory confirmed influenza virus infection between 2009 and 2017 (Adlhoch 2019). In this sample, 83% of patients were ventilated, which is higher than the current rate in Germany (69%). The CFR was 21%, based on a median age of 59 years of admitted patients. Hence, the median age was lower than the median age of death (82 years) currently reported for Germany (Robert Koch Institut 2020). Nevertheless, it needs to be considered that in the SARS-CoV-2 pandemic a portion of the elderly patient population is less likely to be admitted to the ICU and may die, e.g., in nursing homes.

### Willingness to pay

In Germany, the annual treatment costs for new cancer drugs launched between 2011 and 2015 and granted an additional benefit by the German Federal Joint Committee are €65,854, on average (Hammerschmidt, 2017). The average annual costs of comparators are €26,102 (Hammerschmidt, 2017), resulting in incremental costs of €39,751.

Information on the average incremental survival benefit was taken from an analysis of all anticancer drugs launched in Germany between 2011 and 2016 and granted an additional benefit by the German Federal Joint Committee until June 2016. The analysis shows a median incremental survival benefit of 4.7 months or 0.39 years (Storm 2017). This result is similar to what was found in an analysis of 58 anticancer drugs approved in the U.S. between 1995 and 2013, showing an average incremental survival benefit of 0.46 years (Howard 2015). However, in both analyses, the incremental survival benefits are underestimated because they are restricted to the trial period; i.e., they are not extrapolated beyond the trial period (strictly speaking, this is the case only for 47 out of 58 drugs in the study by Howard et al.; see also the Discussion).

Dividing incremental costs by the incremental survival benefit yields €101,493 per life year gained (€39,751/0.39 life years).

## Results

### Health benefits

Based on the independence assumption of mortality rates, a shutdown that is successful in ‘flattening the curve’ is projected to yield a per-capita gain of 0.056 life years (0.67 months) at the time of herd immunity through natural infection (versus no intervention). Multiplying the per-capita gain by the population size and the herd protection threshold results in 3.2 million life years being gained in the German population. At the same time, 1.7 million lives are being saved (calculated by multiplying the difference in CFR compared to no intervention with the population size and the herd protection threshold). These estimates are subject to nonnegligible uncertainty, however. As shown in the sensitivity analysis (see Figure 1), a lower CFR in the population or a higher CFR in the ICU or post discharge reduce the health benefits by approximately one to two thirds. The health benefits of a shutdown also diminish when ICU capacity is exceeded. As shown in Table 2, if the shutdown does not turn out to be successful, exceeding ICU capacity by 50% could reduce the gain in life years by one third.

**Figure 1.**
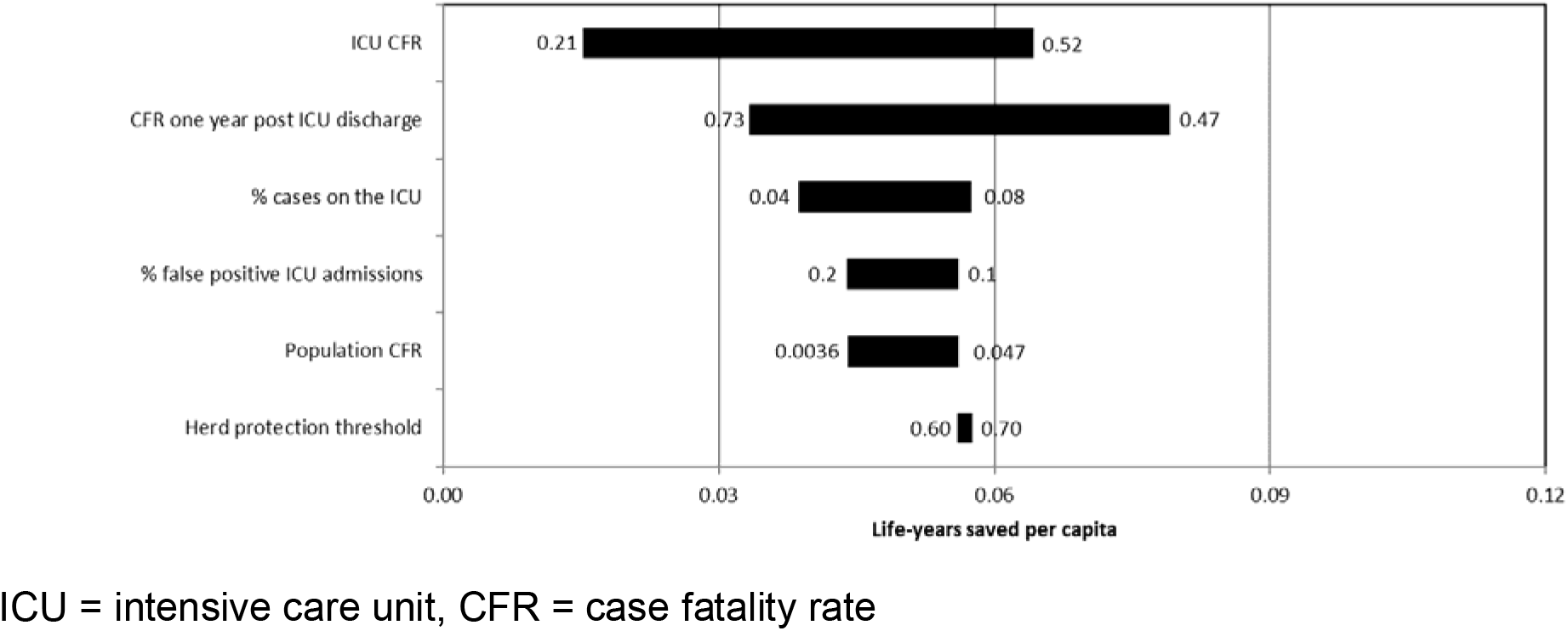
Tornado diagram demonstrating the results of the one-way sensitivity analysis for a shutdown that is successful in ‘flattening the curve’ versus no intervention. Variables are ordered by impact on the number of life years gained per capita. Numbers indicate upper and lower bounds.

**Table 2.** Life years and their monetarized value under different methodological assumptions and strategies.

|  | Independence assumption |  |  | Harvesting assumption |  |  |
| --- | --- | --- | --- | --- | --- | --- |
| Intervention | Per-capita loss of LYs vs. no pandemic | Incremental gain in LYs vs. no intervention | Value of LYs gained (€) | Per-capita loss of LYs vs. no pandemic | Incremental gain in LYs vs. no intervention | Value of LYs gained (€) |
| *'Flattening the curve'* |  |  |  |  |  |  |
| Successful shut-down | 0.418 | 0.056 | 5691 | 0.377 | 0.071 | 7185 |
| ICU capacity exceeded by 50% | 0.440 | 0.035 | 3518 | 0.404 | 0.043 | 4386 |
| ICU capacity exceeded by 100% | 0.458 | 0.016 | 1643 | 0.428 | 0.020 | 2022 |
| ICU capacity exceeded by 200% | 0.469 | 0.005 | 525 | 0.441 | 0.006 | 629 |
| ICU capacity exceeded by 300% | 0.473 | 0.001 | 129 | 0.446 | 0.002 | 159 |
| No intervention | 0.475 | 0 | 0 | 0.447 | 0 | 0 |
| *'Squashing the curve'* |  |  |  |  |  |  |
| Successful shut-down | 0 | 0.475 | 48,160 | 0 | 0.447 | 45,411 |

LY = life year, ICU = intensive care unit

Nevertheless, at the time of herd immunity, even a shutdown that is successful in ‘flattening the curve’ is expected to yield a loss of 0.42 life years per capita compared to the situation before the pandemic. The average number of life years per avoided death is 6.0, which is lower than the average remaining life expectancy in the German population before the pandemic (38.8 years), reflecting the high age of COVID-19 deaths. Based on the harvesting assumption, the average number of life years per avoided death is slightly lower (5.4 years) because the population that dies is assumed to be sicker. On a statistical note, the number of life years per avoided death is equivalent to the change in life years gained by a 1% change in the CFR.

For a newborn, the loss in life expectancy amounts to 0.19 and 0.22 for men and women, respectively. The gain in life years of ‘squashing the curve’ versus no intervention amounts to 0.47 (0.056 + 0.42) life years per capita. By far, the largest driver of this gain is the CFR through the prevalence of undiagnosed cases (see Figure 2).

**Figure 2.**
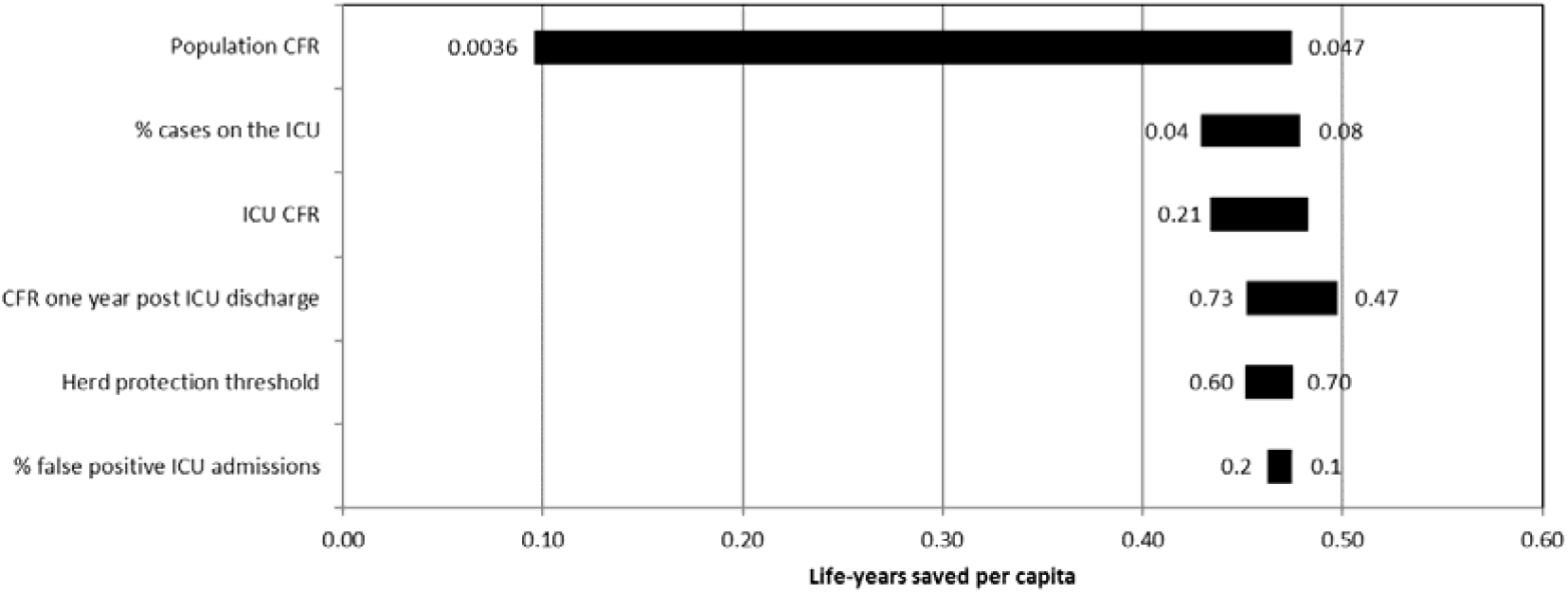
Tornado diagram demonstrating the results of the one-way sensitivity analysis for a shutdown that is successful in ‘squashing the curve’ versus no intervention. Variables are ordered by impact on the number of life years gained per capita. Numbers indicate upper and lower bounds.

### Monetary value

The economic value of a shutdown that is successful in ‘flattening the curve’, based on a per-capita gain of 0.056 life years, is approximately €5691 per capita (see Table 2) or, extrapolated to the total population, 14% of Germany’s GDP in 2019. Using lower and higher estimates based on the sensitivity analysis, the economic value ranges between 4% and 19% of the GDP. The economic value of ‘squashing the curve’ even amounts to 116% of the GDP. It is highly sensitivity to changes in CFR, however: A CFR of 0.36% (Streeck 2020) reduces the economic value to 24%.

### Internal validity

The product of the probabilities of ICU admission and death cannot exceed the CFR in the general population. In fact, given that a portion of deaths occur outside the ICU even with sufficient ICU capacity (e.g., in nursing homes), the product needs to be smaller. This was confirmed (1.9% < 4.7%).

Furthermore, I performed a back-of-the-envelope calculation to verify the health benefits of a shutdown that is successful in ‘squashing the curve’ versus one that is successful in ‘flattening the curve’. Multiplying the average remaining life expectancy in Germany at age 81 (the average age of death) with the increase in CFR compared to the situation before the pandemic and the herd protection threshold yields a number very close to that calculated by the model (0.40 life years versus 0.42 life years).

### External validity

Dividing the number of life years gained by the number of lives saved under ‘flattening the curve’ yields the expected remaining life expectancy of a COVID-19 patient admitted to the ICU (1.9 life years). This number was, by and large, confirmed by observational data from a long-term follow-up study on patients at age 80 and above admitted to an ICU in Norway between 2000 and 2012 (Andersen 2015). In this study, mortality in the ICU and one year after discharge were 24% and 58%, respectively, thus matching well the data informing our model (25% and 55%, respectively). Remaining life expectancy of patients alive at one year after ICU discharge was 5.1 years. When considering the one-year mortality in this study, expected life expectancy of a patient admitted to the ICU is calculated to be 2.1 life years, thus strengthening the validity of our model. A growing life expectancy since the time of conduct of this study can still be accounted for by the harvesting assumption applied in our model, which suggests an expected life expectancy of 2.4 years. Supporting this comparison between simulated and real-world data is the fact that the study by Andersen et al. (2015) was not included in the meta-analysis by Damuth et al. (2015), which informs our model’s post-discharge mortality.

## Discussion

This study uses a life-table model to estimate the impact of a shutdown on lives saved and life years gained in Germany. Although the SARS-CoV-2 pandemic is presumed to be a one-time event, the loss of life years with insufficient ICU capacity is tremendous, yielding a commensurate health gain by a shutdown that is successful in ‘flattening the curve’.

Nevertheless, as herd immunity through natural infection seems unavoidable under ‘flattening the curve’, the loss of life years expressed in terms of the life expectancy of a newborn falls in the range of the annual gain in life expectancy in Germany over the past decade (Federal Office of Statistics 2019). In other words, at the time of herd immunity, the current pandemic will have wiped out the gain in life expectancy obtained from health care and public health interventions in the 12 months preceding the pandemic.

The relatively small loss in life years obtained by assuming that individuals who die from COVID-19 are generally sicker, was indirectly confirmed in another modeling study based on global data (Hanlon 2020). This study showed that explicitly accounting for the various comorbidities associated with COVID-19 “did not drastically impact” the years of life lost due to death from COVID-19.

In addition to comparing the cases of a successful shutdown and no intervention, which forms the base case for the calculation of the maximum monetary value, I analyzed different scenarios with ICU capacity being exceeded to varying degrees. The likelihood of these scenarios depends on the spread of SARS-CoV-2 if the shutdown is not completely effective. Data from 2010/11 indicate that Germany has the highest number of intensive care plus immediate care unit beds on a per-capita basis in Europe (Rhodes 2012). Germany’s leading position in terms of the number of ICU beds was recently confirmed in a report by the Organisation for Economic Co-operation and Development (OECD 2020). ICU capacity will be exceeded, however, when the daily incidence of COVID-19 cases surpasses approximately 16,000 (ZI 2020) (disregarding a potentially lower admission rate due to a high number of undetected COVID-19 cases).

As a word of caution, given the time constraints and the ongoing inflow of new information on the SARS-CoV-2 pandemic while conducting the study and writing this manuscript, which made it pertinent to continuously update the projections, this modeling study has several caveats as discussed below. Given the clear direction of its results, however, it could still provide important guidance for policymakers, which is outlined further below.

First, there are reasons why the base case overestimates the projected health benefits of a shutdown and the corresponding monetary value. Some of these reasons were already described in the sensitivity analysis and include an overestimate of CFR due to a large number of undetected cases. In addition, the study does not consider the deaths and loss of health-related quality of life associated with the shutdown and social distancing, e.g., due to depressive or anxiety disorders, suicides, unemployment, domestic violence, and fewer emergency and physician visits for unrelated medical conditions. Nevertheless, official data on excess mortality in Germany (Federal Office of Statistics 2020) show that both excess mortality and COVID-19 mortality peaked in the first half of April, thus indicating that excess mortality was driven by COVID-19 and not by other causes.

Furthermore, ICU survivors may suffer from a loss of quality of life (Needham 2013). Moreover, unaffected individuals may experience a loss of personal freedom (Abele-Brehm 2020) and autonomy. Finally, if rationing decisions in the ICU disfavor patients with less prospect of survival, the health benefits of a shutdown are reduced (cf. Rühli 2020).

On the other hand, there are reasons to believe that the economic value may be underestimated. First, decreased economic activity can save lives, among other reasons, because it reduces air pollution, traffic accidents (Science Magazine 2020), and accidents on construction sites (Deaton 2020). Second, social distancing may reduce non-COVID-19 flu deaths. Third, reducing the number of deaths prevents grief among caregivers. Forth, COVID-19 containment measures including the shutdown may provide a feeling of security and trust in the government. Fifth, ‘squashing the curve’ might prevent direct (non-)medical costs and indirect costs associated with nonfatal COVID-19 cases. And sixth, the estimate on the one-year mortality of ICU survivors (Damuth 2015) applied in this study represents an overestimate as it includes deaths from diseases unrelated to mechanical ventilation. On the other hand, a potential mortality increase beyond the first year of ICU survival was not modelled. Some of the biases listed in this and the previous paragraph may cancel each other out.

Additional limitations apply to the estimate of the economic value of a life year borrowed from new, innovative cancer drugs. On the one hand, the estimate is too low because the costs of drug-related adverse events and drug-related services are ignored and the costs of cancer treatment are limited to a period of one year. That is, I do not account for the fact that some cancers have a chronic course, thus mandating treatment for more than one year. On the other hand, the estimate is too high because the survival benefit is underestimated because it is confined to the trial period, which typically may not be longer than a year. Again, some of the biases may cancel out.

Furthermore, it may be argued that treatments for conditions more closely related to COVID-19 would provide a more accurate estimate for the economic value. This may include treatments for other viral diseases such as SARS-CoV-1, influenza, or Ebola. Even greater similarity could be obtained by restricting conditions to those that have a similar route of transmission (airborne droplets), CFR, and basic reproduction number (it may reflect a sense of urgency). Nevertheless, the search for a better match may not turn out to be successful. It may also be hampered by the fact that an ideal economic valuation of ‘flattening’ or ‘squashing’ the curve would reflect recently set value-based prices and not the prices of drugs that were launched, for example, more than a decade ago. Using the prices of new, innovative cancer medications as a benchmark also provides an opportunity to check the appropriateness of current cancer prices themselves. If the economic value of ‘flattening’ or ‘squashing’ the curve as determined in this study were considered to be low, then to be consistent, the prices of new cancer drugs would equally deserve a price premium. This implication, however, is contradictory to the present situation, as the pressure to save on health care costs will increase as a consequence of the coronavirus crisis.

The metric of health benefits used in this study, which is the number of life years, may be criticized on ethical grounds. Yet, Emanuel et al. (2020) suggest that “[p]riority for limited resources should aim both at saving the most lives and at maximizing improvements in individuals’ post-treatment length of life […] It is consistent both with utilitarian ethical perspectives that emphasize population outcomes and with nonutilitarian views that emphasize the paramount value of each human life”. Furthermore, the number of life years as an outcome measure may be criticized for lacking a consideration of health-related quality of life. Quality-adjusted life years (QALYs) are able to capture an additional health benefit resulting from the avoidance of non-fatal COVID-19 cases and the associated loss in quality of life under ‘squashing the curve’. On the other hand, QALYs diminish the health benefits obtained from additional life years by accounting for a quality-of-life decrement. As the QALY metric thus discriminates against the elderly and the disabled, it has been considered ethically controversial (Ubel 1999). In the model this caveat applies in particular to ‘flattening the curve’, i.e., when saving patients in the ICU. For these patients there also exists a methodological concern with regard to QALYs when their quality of life is reduced to a degree that they do not want to go on living. This so-called maximum endurable time invalidates the QALY metric (Stalmeier 2001) (as a word of caution, the presence of a maximum endurable time would also question the appropriateness of using life years gained or even lives saved as measures of health benefit but rather for ethical than for methodological reasons). For these reasons, QALYs have not been used so far in Germany for the purpose of reimbursing and pricing new, innovative medicines (cf. IQWiG 2017). As another counterpoint, the public debate on COVID-19 in Germany has mainly focused on mortality as an endpoint and the number of life years lost by the elderly who die from COVID-19. In sum, there is not a straightforward answer to the question of which outcome measure best reflects the value of a successful shutdown. Life years gained may serve as a compromise between the use of unweighted lives saved and QALYs gained.

Given the unprecedented economic value as a share of GDP estimated in this study, the coronavirus crisis also leads to new challenges for economic evaluations in health care and public health. Perhaps the most similar dilemma has arisen surrounding one-off treatments (cures) for genetic disorders. One-off payments for these therapies similarly raise affordability issues even in view of acceptable cost-effectiveness ratios (Towse 2019). Nevertheless, the magnitude of payments required for the COVID-19 shutdown is unparalleled. Hence, while the debate around one-off treatments appears structurally similar, it anticipates only a little of what we are facing.

Even with an acceptable cost-effectiveness ratio of the shutdown, one-time expenditures (corresponding to the economic value of a successful shutdown) would lead to a considerable drop in GDP. In fact, without government intervention, this dip could have major ramifications for financing health care. Considering the maximum economic value of ‘flattening the curve’, a 14% drop in GDP would result in a 16% increase in the portion of total income spent on health care to fund the same type of health care basket (this increase explicitly does not refer to the increase in the contribution rate for SHI, which may be lower than 16%). As the basket does not cover COVID-19, the rise in spending must be viewed independently of the SARS-Cov-2 pandemic and not as an indirect way of supporting the shutdown. If the rise were not acceptable, underfunding of the health care system would result in order to keep spending within an acceptable range. Obviously, this problem is dramatically increased in the case of ‘squashing the curve’.

Hence, there seems to be a tipping point where a drop in income necessitates saving on health expenditures. Defining this tipping point may be starting point for a discussion about the level of GDP decrease that may result in underfunding of the health care system. Of note, mobilizing additional resources for the health care sector through tax financing, national debt, or other means merely shifts the problem of underfunding to employers, other public sectors, or future generations. As a consequence of underfunding, there would be a commensurate loss of lives due to other diseases, thus leading to a zero-sum game. Such opportunity costs are already conceivable at the hospital level, where excess admission for COVID-19 cases strains the health care system and possibly increases mortality from other serious diseases where hospital care is clearly effective (Ioannidis 2020).

Acknowledging this negative feedback loop on the health care system, the way to think about the problem at hand may be in terms of a constrained resource allocation problem, with minimization of life years lost as the objective function and cost-effectiveness ratio, percentage of income spent on health, and ICU capacity as three constraints. Ethical values were already incorporated in the economic value but reappear as an additional constraint when comparing the different exit strategies for lifting social isolation measures. In fact, if agreement on the maximum economic value were reached, the next step would involve assessing the cost-effectiveness of different exit strategies. For the purpose of minimizing the number of life years lost, this would require calculating the degree of excess demand for ICU capacity and the resulting death toll in the absence of intensive care for each exit strategy.

For data collection in the forthcoming months of the crisis, policymakers should pay particular attention to mortality data and the prevalence of undetected cases, as the clinical and economic values forecasted in this study were shown to be particularly sensitive to these data.

## Data Availability

All data analyzed are publicly available.

## Footnotes

1 The number of COVID-19 hospitalizations is less reliable because it includes quarantine as an indication.

2 This framework resembles that of Dworkin’s Theory of Equality (1981), in which an “average member of the community” (ibid., p. 297) behind a ‘thin’ veil of ignorance does not know which handicaps he will eventually develop but does know the probability of becoming handicapped. Using this framework does not necessitate, however, to incorporate all constituents of Dworkin’s Theory of Equality and to accept the resource allocation principles inferred from it.

## Notes

### Competing Interest Statement

The authors have declared no competing interest.

### Funding Statement

This research received no specific grant from any funding agency in the public, commercial or not-for-profit sectors.

### Author Declarations

This is a modelling study using publicly available data.

## References

1. Abele-Brehm A, Dreier H, Fuest C, Grimm V, Kräusslich HG, Krause G, Leonhard M, Lohse AW, Lohse MJ, Mansky T, Peichl A, Schmid RM, Wess G, Woopen C. Die Bekämpfung der Coronavirus-Pandemie tragfähig gestalten: Empfehlungen für eine flexible, risikoadaptierte Strategie. April 2, 2020.

2. Abers MS, Musher DM. Clinical prediction rules in community-acquired pneumonia: lies, damn lies and statistics. QJM. 2014;107(7):595–596.

3. Adlhoch C, Gomes Dias J, Bonmarin I, Hubert B, Larrauri A, Oliva Domínguez JA, Delgado-Sanz C, Brytting M, Carnahan A, Popovici O, Lupulescu E. Determinants of Fatal Outcome in Patients Admitted to Intensive Care Units With Influenza, European Union 2009-2017. Open Forum Infectious Diseases 2019;6(11).

4. Andersen FH, Flaatten H, Klepstad P, Romild U, Kvåle R. Long-term survival and quality of life after intensive care for patients 80 years of age or older. Ann Intensive Care. 2015 Dec;5(1):53.

5. Arentz M, Yim E, Klaff L, Lokhandwala S, Riedo FX, Chong M, Lee M. Characteristics and Outcomes of 21 Critically Ill Patients With COVID-19 in Washington State. JAMA. 2020 Mar 19.

6. Barendregt JJ. The half-cycle correction: banish rather than explain it. Med Decis Mak. 2009;29(4):500–2.

7. Brunner M, Stinner B, Benz SR, Grützmann R. COVID-19-Pandemie: Folgen für die onkologische kolorektale Chirurgie. May 2, 2020. https://www.aerzteblatt.de/nachrichten/112454/COVID-19-Pandemie-Folgen-fuer-die-onkologische-kolorektale-Chirurgie.

8. Bundesministerium für Wirtschaft und Energie. Schwere Rezession durch die Corona-Pandemie. April 29, 2020. https://www.bmwi.de/Redaktion/DE/Schlaglichter-der-Wirtschaftspolitik/2020/05/kapitel-1-1-schwere-rezession-durch-die-corona-pandemie.html.

9. Centre for Evidence-Based Medicine. Global Covid-19 Case Fatality Rates. May 7, 2020. https://www.cebm.net/global-covid-19-case-fatality-rates/.

10. Damuth E, Mitchell JA, Bartock JL, Roberts BW, Trzeciak S. Long-term survival of critically ill patients treated with prolonged mechanical ventilation: a systematic review and meta-analysis. Lancet Respir Med. 2015 Jul;3(7):544-53.

11. Deaton A. Ein freier Markt garantiert keine Gesundheitsversorgung. Die Zeit. April 7, 2020. https://www.zeit.de/wirtschaft/2020-04/angus-deaton-usa-oekonomie-deaths-of-despair.

12. Dworkin R. What is equality? Part 2: Equality of resources. Philosophy & Public Affairs 1981;10:283-345.

13. Emanuel EJ, Persad G, Upshur R, Thome B, Parker M, Glickman A, Zhang C, Boyle C, Smith M, Phillips JP. Fair Allocation of Scarce Medical Resources in the Time of Covid-19. N Engl J Med. 2020 Mar 23.

14. Federal Office of Statistics. Allgemeine Sterbetafel. Wiesbaden: Federal Office of Statistics; 2019.

15. Federal Office of Statistics. Lebenserwartung steigt nur noch langsam. Pressemitteilung Nr. 427 vom 5. November 2019. Wiesbaden: Federal Office of Statistics; 2019.

16. Federal Office of Statistics. Vorausberechneter Bevölkerungsstand. Moderate Entwicklung der Geburtenhäufigkeit, Lebenserwartung und Wanderung (G2L2W2). Wiesbaden: Federal Office of Statistics; 2020.

17. Federal Office of Statistics. Rund 500 000 Krankenhausbetten im Jahr 2017. Pressemitteilung Nr. N 011 vom 13. März 2020. https://www.destatis.de/DE/Presse/Pressemitteilungen/2020/03/PD20_N011_231.html;jsessionid=FF415DD7AB04D7AED61779FAB892D46F.internet8712 (accessed on April 1, 2020).

18. Federal Office of Statistics. Sonderauswertung zu Sterbefallzahlen des Jahres 2020. Juni 19, 2020. https://www.destatis.de/DE/Themen/Gesellschaft-Umwelt/Bevoelkerung/Sterbefaelle-Lebenserwartung/sterbefallzahlen.html.

19. Financial Times. UK strategy likely to cause 35,000-70,000 excess deaths, says study. March 22, 2020. https://www.ft.com/content/f3796baf-e4f0-4862-8887-d09c7f706553.

20. Gandjour A. A proportional rule for setting reimbursement prices of new drugs and its mathematical consistency. BMC Health Services Research 2020;20:240.

21. Garrison LP Jr, Mansley EC, Abbott TA 3rd, Bresnahan BW, Hay JW, Smeeding J. Good research practices for measuring drug costs in cost-effectiveness analyses: a societal perspective: the ISPOR Drug Cost Task Force report--Part II. Value Health. 2010 Jan-Feb;13(1):8-13.

22. Hammerschmidt T. Analyse der AMNOG-Erstattungsbeträge im europäischen Preisumfeld. Gesundh Ökon Qual Manag. 2017;22(1):43–53.

23. Hanlon P, Chadwick F, Shah A, et al. COVID-19 – exploring the implications of long-term condition type and extent of multimorbidity on years of life lost: a modelling study [version 1; peer review: awaiting peer review] Wellcome Open Research 2020;5:75.

24. Howard DH, Bach PB, Berndt ER, Conti RM. Pricing in the market for anticancer drugs. J Econ Perspect. 2015;29(1):139–162.

25. Institut für Qualität und Wirtschaftlichkeit im Gesundheitswesen. General methods: version 5.0. Cologne, Germany; 2017.

26. Ioannidis JPA. Coronavirus disease 2019: the harms of exaggerated information and non-evidence-based measures. Eur J Clin Invest. 2020 Mar 23:e13223.

27. Kupferschmidt K, Cohen J. Race to find COVID-19 treatments accelerates. Science 27 Mar 2020; 367(6485):1412-1413.

28. Kwok KO, Lai F, Wei WI, Wong SYS, Tang J. Herd immunity - estimating the level required to halt the COVID-19 epidemics in affected countries. J Infect. 2020 Jun; 80(6):e32–e33.

29. Needham DM, Dinglas VD, Bienvenu OJ, Colantuoni E, Wozniak AW, Rice TW, Hopkins RO; NIH NHLBI ARDS Network. One year outcomes in patients with acute lung injury randomised to initial trophic or full enteral feeding: prospective follow-up of EDEN randomised trial. BMJ. 2013 Mar 19;346:f1532.

30. Organisation for Economic Co-operation and Development. Beyond Containment: Health systems responses to COVID-19 in the OECD. Paris: OECD; 2020.

31. Rhodes A, Ferdinande P, Flaatten H, Guidet B, Metnitz PG, Moreno RP. The variability of critical care bed numbers in Europe. Intensive Care Med. 2012 Oct;38(10):1647-53.

32. Robert Koch Institut. Bericht zum Krebsgeschehen in Deutschland 2016. Berlin: Robert Koch Institut; 2016.

33. Robert Koch Institut. https://www.rki.de/DE/Content/InfAZ/N/Neuartiges_Coronavirus/Situationsberichte/Gesamt.html. June 15, 2020.

34. Rühli L. Wann schadet der Corona-Lockdown mehr, als er nützt: Ethik und Ökonomie als zwei Seiten einer Medaille. March 30, 2020. https://www.avenir-suisse.ch/wann-schadet-der-lockdown-mehr-als-er-nuetzt/.

35. Science Magazine. Can you put a price on COVID-19 options? Experts weigh lives versus economics. March 31, 2020. https://www.sciencemag.org/news/2020/03/modelers-weigh-value-lives-and-lockdown-costs-put-price-covid-19#.

36. Stalmeier PF, Chapman GB, de Boer AG, van Lanschot JJ. A fallacy of the multiplicative QALY model for low-quality weights in students and patients judging hypothetical health states. Int J Technol Assess Health Care. 2001;17(4):488–496.

37. Storm A, Greiner W, Witte J. AMNOG-Report 2017: Nutzenbewertung von Arzneimitteln in Deutschland. DAK-Gesundheit; 2017.

38. Streeck H, Schulte B, Kuemmerer B, et al. Infection fatality rate of SARS-CoV-2 infection in a German community with a super-spreading event. medRxiv 2020.05.04.20090076; doi: https://doi.org/10.1101/2020.05.04.20090076.

39. Sud A, Jones M, Broggio J, Loveday C, Torr B, Garrett A, Nicol DL, Jhanji S, Boyce SA, Gronthoud F, Ward P, Handy JM, Yousaf N, Larkin J, Suh YE, Scott S, Pharoah PDP, Swanton C, Abbosh C, Williams M, Lyratzopoulos G, Houlston R, Turnbull C. Collateral damage: the impact on outcomes from cancer surgery of the COVID-19 pandemic. Ann Oncol. 2020 May 16:S0923-7534(20)39825-2.

40. Towse A, Fenwick E. Uncertainty and Cures: Discontinuation, Irreversibility, and Outcomes-Based Payments: What Is Different About a One-Off Treatment? Value Health. 2019 Jun;22(6):677-683.

41. Ubel PA, Richardson J, Prades JL. Life-saving treatments and disabilities. Are all QALYs created equal?. Int J Technol Assess Health Care. 1999;15(4):738–748.

42. Wall Street Journal. Human Testing Begins Earlier Than Expected For U.S. Coronavirus Vaccine. March 28, 2020. https://www.wsj.com/articles/u-s-coronavirus-vaccine-study-begins-earlier-than-expected-11584379352

43. World Health Organization. What is a pandemic? https://www.who.int/csr/disease/swineflu/frequently_asked_questions/pandemic/en/

44. Wu Z, McGoogan JM. Characteristics of and Important Lessons From the Coronavirus Disease 2019 (COVID-19) Outbreak in China: Summary of a Report of 72-314 Cases From the Chinese Center for Disease Control and Prevention. JAMA. 2020 Feb 24.

45. Zentralinstitut für die kassenärztliche Versorgung in Deutschland. Kennzahlen zum Management der COVID-19-Pandemie durch die Bundesländer. May 12, 2020. https://www.zi.de/fileadmin/images/content/PMs/Statement_COVID19_Belastungsgerenze_2020-05-12.pdf.

